# Attributing heatwave mortality to human-induced climate change in Greece: a case-crossover and attribution analysis for 2000-2019

**DOI:** 10.64898/2026.03.25.26349303

**Authors:** Di Xi, Dimitris Evangelopoulos, Clair Barnes, Evangelos Chandakas, Constantine Vardavas, Paraskevi Katsaounou, Paolo Vineis, Filippos T Filippidis, Garyfallos Konstantinoudis

## Abstract

**Background:** Heatwaves increasingly threaten public health in the Mediterranean region, and Greece is among the hardest hit countries. Yet evidence on long-term adaptation, spatial vulnerability, and the contribution of human-induced climate change to heatwave-related mortality in Greece remains limited.

**Methods:** We analysed 2,144,957 all-cause deaths in Greece (2000–2019) using a time-stratified case-crossover design. We derived population-weighted daily maximum temperatures at NUTS3 level from ERA5 reanalysis and WorldPop. We applied six heatwave definitions (HD1–HD6) varying by duration (≥2 or ≥3 days) and thresholds (90th, 95th, 99th percentiles).

We fitted Bayesian hierarchical Poisson models to estimate heatwave-mortality associations varying by space and time. We additionally adjusted for relative humidity and national. We then combined these estimates with probabilistic climate-attribution methods to quantify the number and proportion of heatwave-related deaths attributable to human-induced climate change.

**Results:** Heatwaves raised mortality consistently, with relative risks from 1.08 (95% CrI (Credible Interval): 1.07–1.09; HD1) to 1.15 (1.11–1.20; HD6). Risks increased with heatwave intensity and duration and peaked among females and adults aged ≥85 years. We did not detect a consistent temporal decline in risk or marked spatial heterogeneity. Human-induced climate accounted for 51-94% of heatwave-related deaths across definitions. The proportion attributable to climate change rose over time.

**Conclusions:** Heatwaves already impose a major mortality burden in Greece, with more than half driven by anthropogenic climate change and little evidence of population-level adaptation. These findings call for rapid emissions reductions and targeted adaptation, including stronger heat-health warning systems and protection of vulnerable groups.

**What our research question was:** What are the spatiotemporal patterns of heatwave-related mortality in Greece, and what is the contribution of human-induced climate change to these patterns?

**What we found:** Heatwaves were associated with an increased risk of mortality, with no clear evidence of spatial heterogeneity or temporal adaptation. Climate change substantially amplified these effects, accounting for more than half of heatwave-associated deaths.

**Why it is important:** These findings indicate that current measures are insufficient to reduce the health burden of heatwaves, highlighting the urgent need for rapid emissions reductions and targeted adaptation strategies, including stronger heat–health warning systems and improved protection of vulnerable populations.

## Introduction

Temperatures around the globe are increasing due to anthropogenic emissions of greenhouse gases (1). Record-breaking temperatures observed in Europe during the 2022 summer were associated with more than 60,000 heat-related deaths (2). Nearly half of these deaths are estimated to be attributable to human-induced climate change, with Southern European countries being disproportionately affected (3). Focusing specifically on deaths attributable to climate change can strengthen climate risk communication and enhance “subjective attribution”, the public’s growing recognition that extreme events are linked to climate change, which is a key driver of support for climate mitigation policies (4). Greece is among the countries in Europe most affected by increasing warming; however, there is still a lack of nationwide studies quantifying the heat-related health impacts of anthropogenic climate change.

Heat exposure can be defined as ambient exposure to higher temperatures or heatwaves. Heatwaves, defined as prolonged periods of unusually high temperatures above certain thresholds, have been reported to be associated with increased morbidity and mortality globally (2,5–12). Focusing on heatwaves rather than ambient temperature offers clearer cut-off points to activate targeted public health actions, such as health-heat early warnings (13). The first ever health heat attribution study examined the extreme European heatwaves of summer 2003 and showed that anthropogenic climate change increased the risk of heat-related mortality by ∼70% in Central Paris and by ∼20% in London (14). A global study reported that more than half of heatwave-related deaths in 2023 were attributable to human-induced climate change (15). The first ever rapid health attribution study was focused on 12 European cities during a 10-day heat period in summer 2025 and reported that human-induced climate change has tripled the heat-related mortality toll (16). However, all these studies rely on a single estimate of the effect of heat on mortality and do not account for possible changes over time due to adaptation. In addition, most have focused on specific events, and none has examined long-term trends in heat-related mortality attributable to climate change.

Using nationwide data in Greece during 2000-2019, we investigated the effect of heatwaves on all-cause mortality using six heatwave definitions. We examined temporal adaptation and spatial vulnerabilities across the different heatwave definitions, sex and age groups. We then simulated counterfactual temperature time series assuming no anthropogenic climate change and compared with the factual temperature time series to isolate the footprint of human-induced climate change on heat-related mortality in Greece. Finally, we analysed temporal trends in heat-related mortality driven by climate change over 2000-2019.

## Methods

### Data

We retrieved individual level all-cause mortality for individuals aged 65 and above during 2000-2019 in Greece from the Hellenic Statistical Authority (17). Data included information about age, sex, region of residence classified according to the Nomenclature of Territorial Units for Statistics at level 3 (NUTS3) and the date of death (18). We grouped the data by age (65-74, 75-84 and >85) and sex (males and females). The selection of the age subgroups was in line with the literature (19–21).

### Exposure and confounders

We retrieved daily maximum temperature for summer months (June-August) during 2000-2019 at approximately 10km resolution from ERA5-land reanalysis dataset (22). For each NUTS3 region, we calculated its maximum daily population-weighted temperatures using population data from WorldPop at 1 km geographical resolution (www.worldpop.org), see Supplementary Methods Section 1.We also obtained daily mean relative humidity from the ERA5-land reanalysis dataset and calculated the population-weighted estimate in similar way as for temperature (22).

### Heatwave definition

Heatwave is defined as a prolonged period of unusual high temperatures exceeding certain threshold within a given region, typically lasting for a duration of several consecutive days. The temperature thresholds in our analysis were region- and year-specific, being estimated using the 90^th^, 95^th^ and 99^th^ percentiles of the yearly distribution of maximum temperature for each NUTS3 region. We use six heatwave definitions (HD1-HD6), to capture different combinations of heat intensity and duration. Specifically, HD1 refers to periods of at least two consecutive days with maximum temperature above the 90th percentile, while HD2 refers to at least three consecutive days above the same threshold.

Similarly, HD3 and HD4 correspond to at least two and three consecutive days above the 95^th^ percentile, respectively, and HD5 and HD6 represent at least two and three consecutive days above the 99th percentile. We also considered a 3-day lag to account for prolonged effects, whereby a day was classified as a heatwave day if a heatwave had occurred within the preceding three days.

### Counterfactual temperatures

The change in heatwave temperatures in each region associated with each degree of global warming was estimated using the probabilistic attribution protocol established by World Weather Attribution (23,24). Briefly, a nonstationary Generalised Extreme Value distribution is fitted to the time series of maxima, in which the location parameter of the distribution is assumed to shift linearly with global mean surface temperature. The output of this model is how much warmer each year was than a preindustrial climate with no or minimal human-caused warming. We repeated this procedure for 2 observation-based and 72 regional climate model datasets and synthesised the results. These estimates were used to constructed counterfactual temperature in a world without human-including warming, see Supplementary Methods Section 2.

### Study design

We used a time-stratified case-crossover design, commonly used for analysing the effect of transient exposures(25). The temperature on the day of mortality (event day) is compared with the temperature on non-event days. In the case-crossover design, a case serves as its own control, thus, this design automatically controls for factors that do not vary or vary slowly over time, such as sex or deprivation. We selected non-event days on the same day of week and calendar month as the event day to avoid the overlap bias (26).

### Epidemiological model

We modelled the effect of heatwave on mortality by specifying Bayesian hierarchical conditional Poisson models, with a fixed effect on the event/non-event day grouping. This has been found to provide an established flexible alternative to conditional logistic regression (27). We used conditional autoregressive priors to allow the heatwave effects to vary in space and time capturing regional and temporal heterogeneity. Effects were adjusted for relative humidity and national holidays. We have run the models for the different age-sex groups separately. We report median and 95% Credible Intervals (CrI) of the posterior of the relative heatwave mortality risk by age, sex, heatwave definition, time and space (Supplementary Methods Section 3).

### Death attribution

We estimated the nationwide number of deaths attributable (AN) to each heatwave definitions during the study period, based on factual and counterfactual temperatures and stratified by age and sex group. AN were estimated by combining the heatwave relative risks with the number of heatwave days under factual and counterfactual temperature scenarios, enabling quantification of mortality attributable to anthropogenic climate change, Supplementary Methods Section 4. All analyses were conducted using the Integrated Nested Laplace Approximation (INLA) (28,29).

## Results

### Descriptive analysis

We observed a total of 2,144,957 deaths in Greece during 2000-2019, of which 49.1% (1,052,792) were among females (Table 1). The number of deaths observed during heatwaves declined as the heatwave definitions became more extreme (Table 1). For example, there were 281,503 deaths under the least extreme definition (HD1), compared to 7,780 deaths under the most extreme definition (HD6). Descriptive statistics of the exposure are included in the Supplementary Table 1, Online Supplemental Figure 1 and 2. Overall, we did not observe any consistent spatial or temporal patterns in the heatwave exposure, reflecting the choice of the percentiles in the heatwave definition.

**Table 1.**
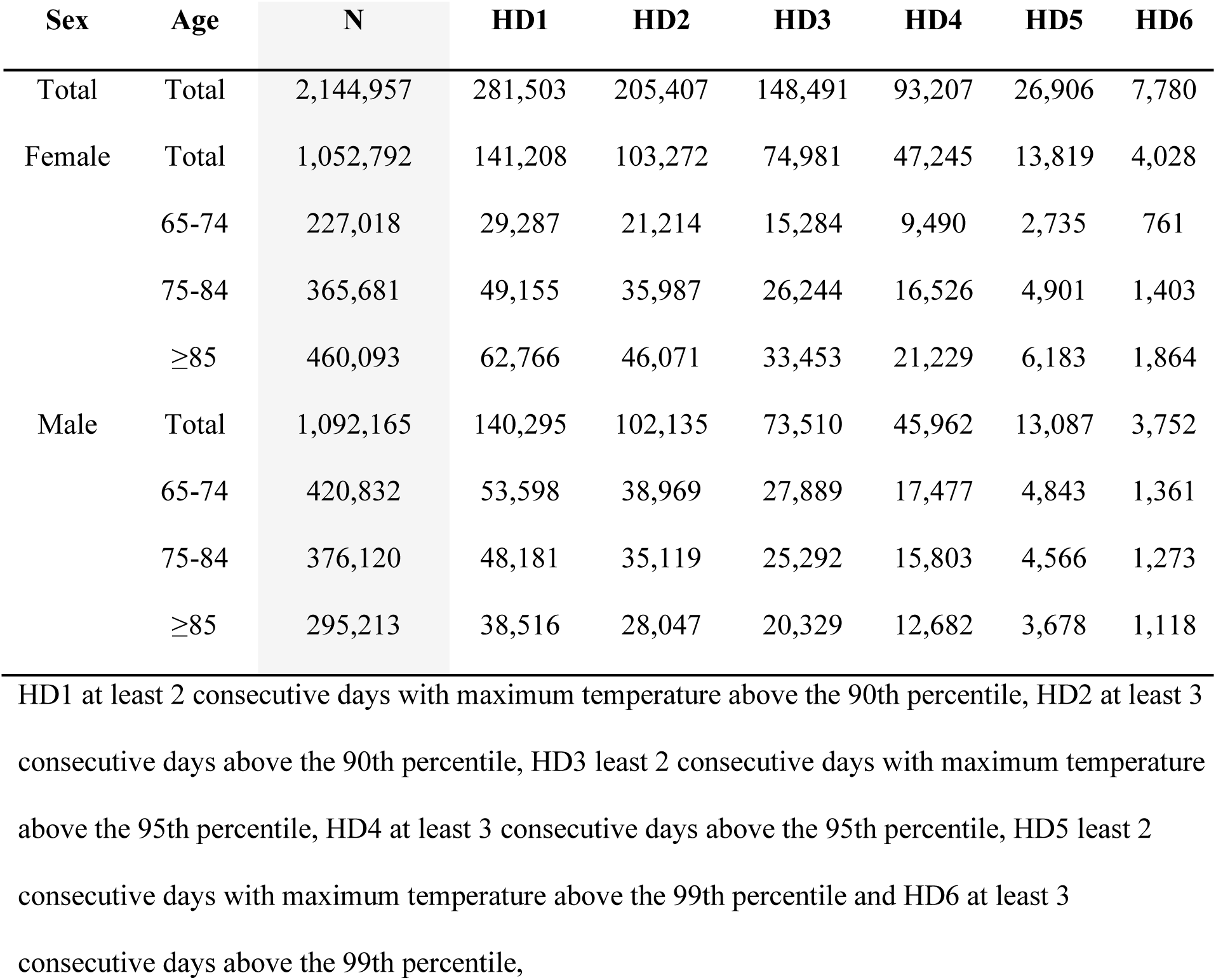
Number of all-cause deaths across age and sex group and the six different heatwave definitions in Greece from 2000 to 2019.

### Main heatwave effect

Across the study period and geographical regions, we observed a strong association between heatwaves and all-cause mortality. The relative risk (RR) for mortality on heatwave days versus non-heatwave days ranged from 1.11 (95% CrI: 1.11-1.12, HD1) to 1.25 (95% CrI: 1.21-1.28, HD6) in unadjusted models (not accounting for relative humidity and national holidays), and 1.08 (95% CrI: 1.07-1.09, HD1) to 1.15 (95% CrI: 1.11-1.20, HD6) in fully adjusted models (Figures 1, Supplementary table 2). The effect of heatwaves on mortality increased with increasing intensity and duration (Figure 1). However, for more extreme heatwave definitions, the number of heatwave events is lower, leading to greater uncertainty in the estimated effects (Figure 1, Supplementary Table 2).

**Fig 1.**
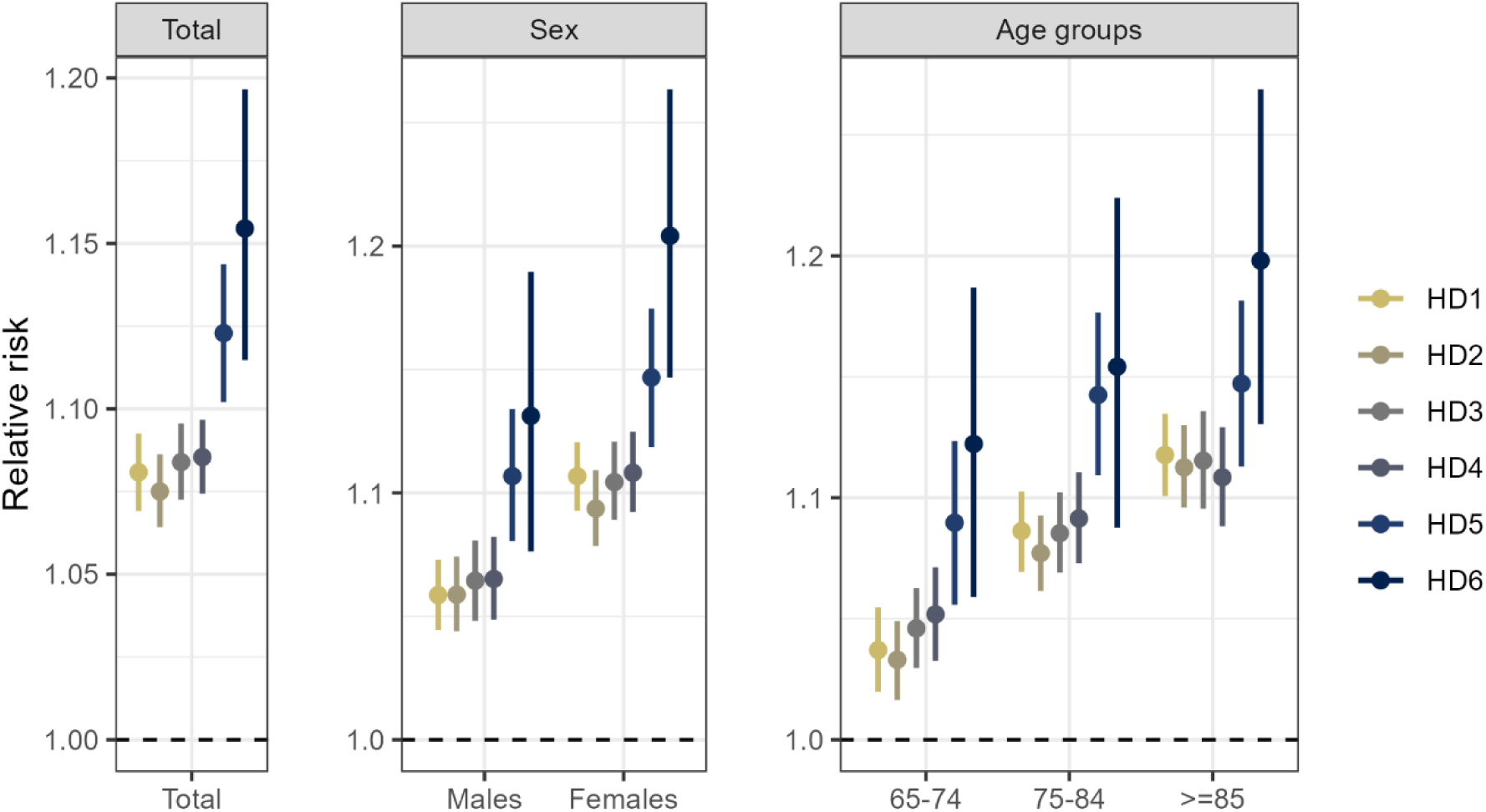
Median and 95% credible intervals of the posterior distribution of the relative heatwave risk from the fully adjusted model across the 6 heatwave definitions. HD1 at least 2 consecutive days with maximum temperature above the 90th percentile, HD2 at least 3 consecutive days above the 90th percentile, HD3 least 2 consecutive days with maximum temperature above the 95th percentile, HD4 at least 3 consecutive days above the 95th percentile, HD5 least 2 consecutive days with maximum temperature above the 99th percentile and HD6 at least 3 consecutive days above the 99th percentile.

Across the different sex and age groups, overall females and older populations were more vulnerable, with the highest RR among females 75-84 years old: 1.24 (95% CrI: 1.15-1.33) (Supplementary Table 2).

### Spatiotemporal variation of the heatwave effect

Figure 2 shows the spatiotemporal trends of the heatwave effect across the six considered definitions. Panel A of Figure 2 presents the percentage change in the heatwave effect over time. Overall, there are no consistent temporal trends in the heatwave effects across the different definitions. The smallest heatwave effect was observed in 2019, and this finding was consistent across all heatwave definitions (Panel A, Figure 2). Panel B of Figure 2 displays the posterior probability that the area-specific heatwave effect is larger than the overall nationwide effect across the different heatwave definitions. We found no strong evidence (i.e., posterior probability always smaller than 0.95) of spatial heterogeneity in the heatwave effect, with the highest posterior probability of a spatial deviation from the overall effect being 0.93 for HD1 in the Dodecanese Islands (Panel B, Figure 2).

**Fig 2.**
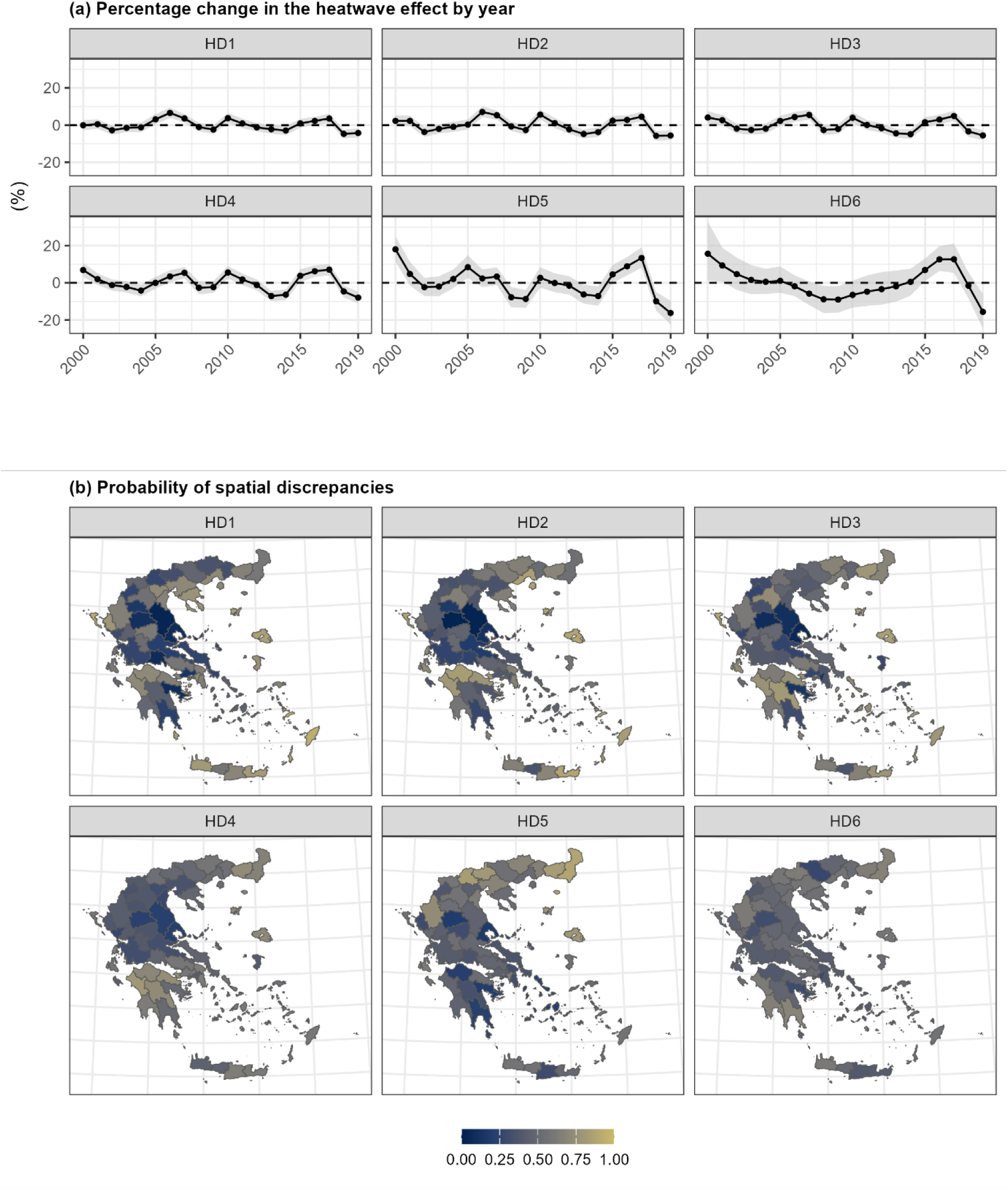
Spatiotemporal variation of the heatwave effects across the 6 heatwave definitions. Panel A shows the median percentage change in the heatwave effect across the years, whereas panel B the posterior probability that the area-specific heatwave effect is larger than the overall nationwide effect across the different heatwave definitions. Both effects were based on the fully adjusted models. HD1 at least 2 consecutive days with maximum temperature above the 90th percentile, HD2 at least 3 consecutive days above the 90th percentile, HD3 least 2 consecutive days with maximum temperature above the 95th percentile, HD4 at least 3 consecutive days above the 95th percentile, HD5 least 2 consecutive days with maximum temperature above the 99th percentile and HD6 at least 3 consecutive days above the 99th percentile,

### The impact of human-induced climate change

Figure 3 shows the number of heatwave days in the factual and counterfactual scenarios across the different heatwave definitions. Overall, the number of heatwave days has increased over time across all heatwave definitions. Table 2 presents the number of additional heatwave deaths and the proportion of deaths attributed to human-induced climate change. For the milder heatwave definitions, approximately half of the heatwave-related deaths during the study period were attributable to human-induced climate change. For example, under HD1, there were 19,711 (95% CrI: 16,763 - 22,034) expected heatwave-related deaths, of which 51% (95% CrI: 43 - 56%) were attributable to climate change. In contrast, for the stricter definitions, nearly all heatwave-related deaths could be linked to anthropogenic climate change. For instance, under HD6, there were 1,207 (95% CrI: 1,015 - 1,412) heatwave-related deaths, 94% (95% CrI: 90 - 96%) of which were attributable to climate change. Last, we calculated the annual number of expected heatwave-attributable deaths and the corresponding proportions attributable to climate change by year. The number of deaths showed no consistent temporal pattern, whereas the proportion increased over time (Figure 4).

**Fig 3.**
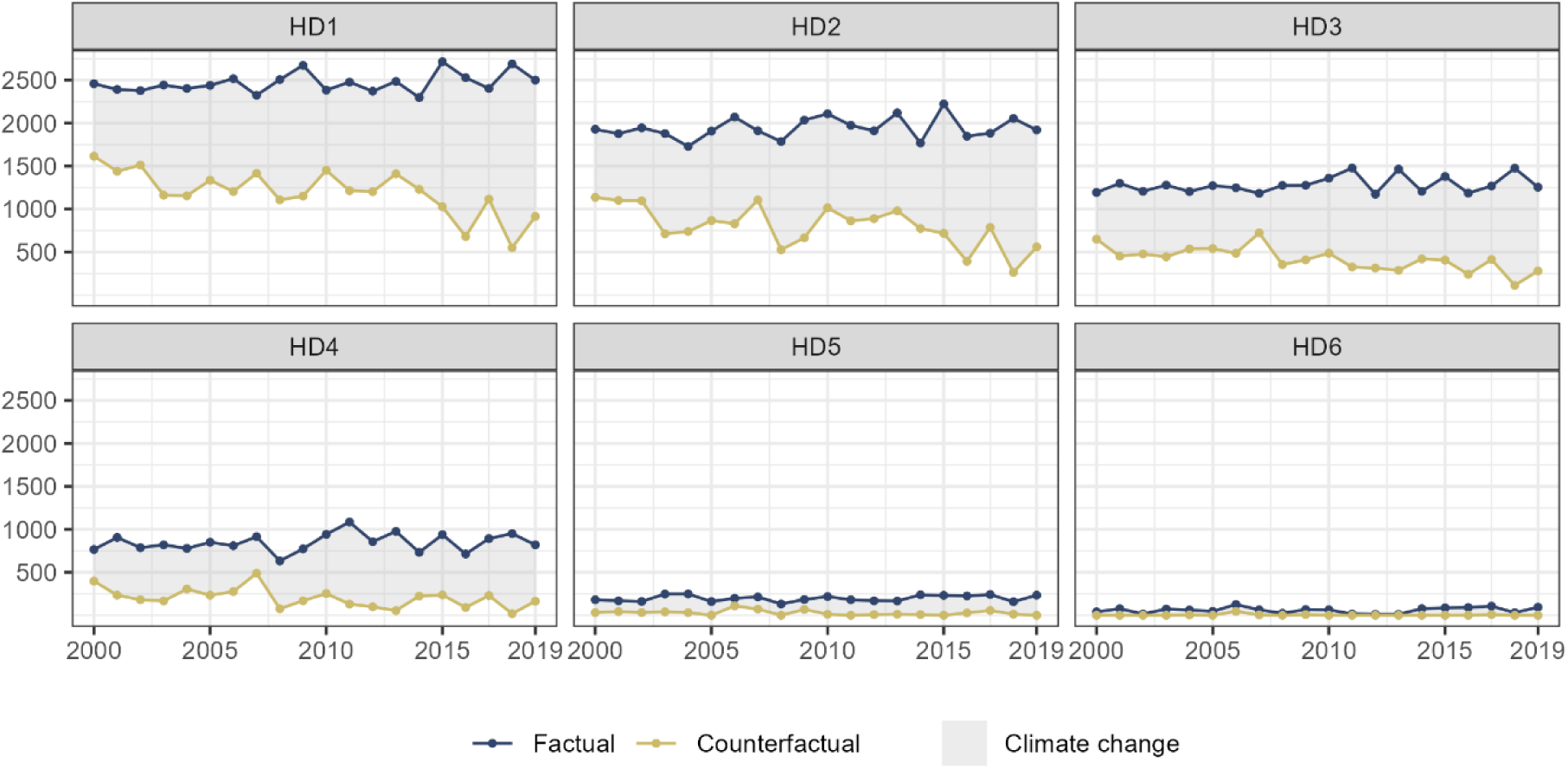
Numbers of heatwave days (y-axis) in factual and counterfactual scenarios over the study period across (x-axis) the 6 heatwave definitions. HD1 at least 2 consecutive days with maximum temperature above the 90th percentile, HD2 at least 3 consecutive days above the 90th percentile, HD3 least 2 consecutive days with maximum temperature above the 95th percentile, HD4 at least 3 consecutive days above the 95th percentile, HD5 least 2 consecutive days with maximum temperature above the 99th percentile and HD6 at least 3 consecutive days above the 99th percentile,

**Fig 4.**
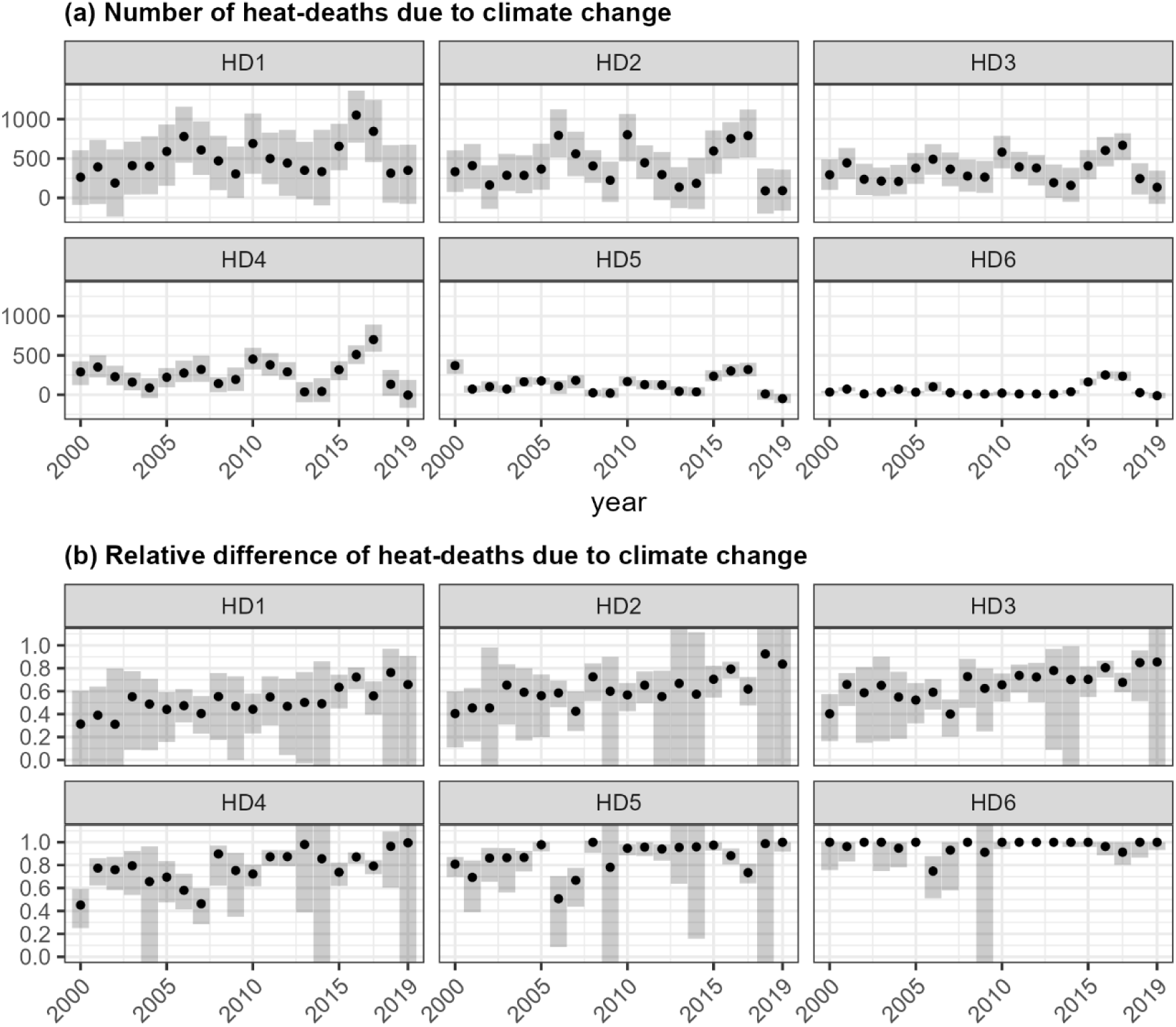
Median and 95% credible intervals of the number of heat-deaths and relative difference of deaths due to human-induced climate change across the study period. HD1 at least 2 consecutive days with maximum temperature above the 90th percentile, HD2 at least 3 consecutive days above the 90th percentile, HD3 least 2 consecutive days with maximum temperature above the 95th percentile, HD4 at least 3 consecutive days above the 95th percentile, HD5 least 2 consecutive days with maximum temperature above the 99th percentile and HD6 at least 3 consecutive days above the 99th percentile,

**Table 2.**
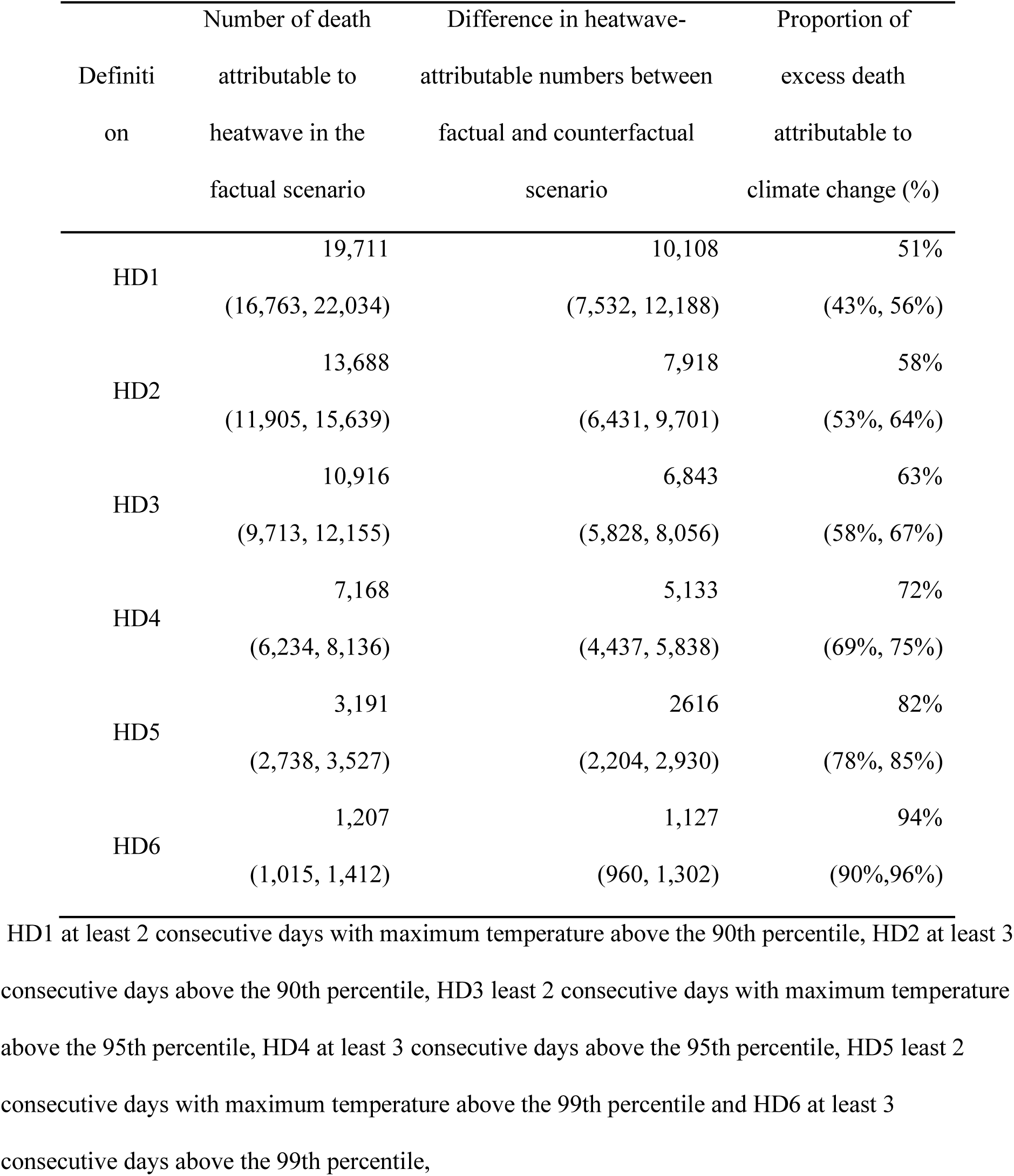
Median and 95% credible intervals of number of deaths attributable to heatwaves in the factual scenario, number of heatwave deaths and the proportion of deaths attributed to human-induced climate change. The results are calculated using relative risks from the fully adjusted models.

## Discussion

In this first nationwide spatiotemporal study in Greece during 2000-2019, we investigated the effect of heatwaves on all-cause mortality using different heatwave definitions. We examined temporal adaptation and spatial vulnerabilities, as well as the effect modification by age and sex. We found that heatwaves were associated with increased all-cause mortality, with higher-intensity heatwaves associated with greater risk. Older individuals and females were more vulnerable. We found weak evidence of spatiotemporal variation of the heatwave effect across the different heatwave definitions. We observed that human-induced climate change has increased the number of days exceeding the high temperature percentiles of the climatology and the associated number of heatwave-related deaths. Around half of heatwave-related deaths were attributable to human-induced climate change for less stringent heatwaves, while nearly all deaths could be linked to climate change under the stricter definitions.

The results of this study identified population groups that were more impacted than others. This demographic pattern aligns with previous heat-mortality studies, which consistently report elevated risks among older populations (2,5,7,8,12,20) and females (2,5,8,12,20,30) across different climatic and geographic contexts. Several biological and physiological mechanisms may explain this heightened susceptibility (31,32). Older adults often have reduced thermoregulatory capacity, diminished cardiovascular reserve, and a higher prevalence of chronic conditions that impair the ability of heat dissipation. Females may experience greater vulnerability due to differences in body composition, hormonal regulation, and higher rates of heat-sensitive comorbidities (33,34).

In our analysis, we did not detect substantial spatial variation in heatwave-related mortality risk across regions. Previous studies focusing on mortality or hospital admissions and ambient temperature have reported some evidence of spatial effect modification that can be partly explained by environmental covariates such as green space and urbanicity (35–38). The lack of spatial heterogeneity in our study could reflect relatively homogeneous climatic conditions across much of Greece during extreme heat events, nationwide exposure patterns, or similarities in demographic, behavioural (e.g. smoking and physical activity) and health system characteristics that influence vulnerability. It may also be partly due to limitations in statistical power to detect small-scale geographic differences.

Our results provide weak evidence of population-wide adaptation to heatwaves in Greece over the study period. This limited adaptation contrasts with findings from other Mediterranean European countries, such as Spain (39–41), where previous studies have reported a reduction in heat-related mortality risk and attributable deaths over time. A multi-country study also found a decrease in heat-related mortality impacts over the past decades, beyond what would be expected from adaptation alone given the observed warming in most of the studied countries (42). However, there was only weak evidence of adaptation in Australia and Brazil (42). A study using partly overlapping data focusing on Thessaloniki (1999–2018), Greece’s second-largest city, and Athens (1999–2019), the capital, reported steeper temperature-mortality curves in Athens during 2011-2019 compared to 1999-2007, while curves in Thessaloniki remained largely unchanged (43).

This discrepancy may reflect methodological differences from previous studies, including the choice of exposure metric (heatwaves versus ambient temperature), the selection of percentiles and duration criteria, the consideration of lag periods, as well as differences in study period, study region, and statistical model. The consistency of our results with the previous Greek study further supports our conclusion of limited adaptation to heat in Greece. In contrast, the evidence of adaptation reported in other Mediterranean countries, such as Italy and Spain (42,44), may reflect the greater effectiveness of their heat-health warning systems, better hospital preparedness, and more comprehensive public health interventions. In Greece, the apparent lack of adaptation could be linked to less efficient heat-health alert systems, limited investment in green space and urban cooling strategies, and insufficient integration of air pollution reduction policies with heat mitigation plans. Strengthening these areas could enhance resilience to rising temperatures and reduce the future health burden of heat.

Our study is consistent with previous studies quantifying the impact of human-induced climate change on heat-related mortality, especially under milder heatwave definitions. For example, under HD1 and HD2, 51% (95% CrI: 43%-56%) and 58% (95% CrI: 53%-64%) of heat-related death can be attributable to climate change. This is in line with a global study reporting that 54.3% of heatwave-related deaths were attributable to human-induced climate change (15), as well as a European study in the summer of 2022, which found that 56% of observed heat-related deaths could be attributed to climate change(2). Similarly, a study in Switzerland estimated this proportion at 60% (45).

Furthermore, our study reported most heat-related deaths can be linked to climate change under the stricter definitions. For instance, under HD4, 72% (95% CrI: 69%-75%) of heat-related death can be attributable to climate change, and under HD6, this proportion reached 94% (95% CrI: 90%-96%). This number is consistent with a recent rapid health attribution study focusing on 854 cities in Europe, reporting 70% of the expected heat-related mortality to be attributed to climate change (46). These patterns show that as heatwaves become more intense, a growing proportion of the resulting mortality is attributable to human-induced climate change, underscoring the heightened public-health risk under more extreme warming.

The main strength of this study is the availability of a large nationwide dataset with granular spatial information in Greece during 2000-2019 making the results generalisable to both urban and rural settings. The availability of individual data minimises ecological bias (47). We proposed a Bayesian spatiotemporal framework to estimate the effect of heatwaves in space and time by borrowing information from spatial and temporal neighbours. This aims to smooth the heatwave effects towards local and global means and accounts for data sparsity. Nevertheless, as the heatwaves are relatively rare events, this study had limited power to detect changes in space for more extreme heatwave definitions.

Our study has some limitations. Residential temperature does not reflect the actual temperature exposure of an individual, as individuals are exposed to different temperatures during the day. In addition, the outdoor temperature, does not always reflect the actual temperature exposure in indoor spaces. Nevertheless, in line with most of the studies in this field and given the lack of more precise individual exposure data, we used residential temperature outdoors as a proxy for the individual exposure. Our analysis lacked information on additional individual-level factors that could influence vulnerability, including access to air conditioning, smoking status and medication use. Due to the lack of daily air pollution data on high geographical resolution, we could not account for the effect of air pollution, nevertheless, we assumed that it is a potential mediator of the observed effect(35).

Additionally, in our time-stratified case-crossover designs, some control days may occur after the death event. However, this sampling is not expected to introduce bias because environmental exposures are exogenous to individual mortality.

## Conclusion

Our findings highlight the substantial health burden attributable to heatwaves in Greece, particularly among older adults and females. As climate change increases the frequency and intensity of extreme heat events, proactive adaptation measures, such as heat–health warning systems, community outreach, and tailored guidance for vulnerable populations, will be critical to reducing mortality. The absence of a temporal decline in heatwave-related mortality risk suggests limited adaptation or ineffective implementation of existing measures. Our counterfactual analysis indicates that a significant proportion of heatwave-related deaths are attributable to human-induced climate change, highlighting the importance of both mitigation policies to curb greenhouse gas emissions and adaptation strategies to protect public health. Protecting vulnerable populations through early warning systems, public health preparedness, and community-based interventions, together with mitigation actions such as decarbonizing the energy sector and expanding urban green spaces, will be essential to reduce future heatwave-related deaths in Greece.

## Contributors

G.K. conceived the study. G.K., D.E. and F.F. supervised the study. G.K., D.E. and F.F developed the initial study protocol. G.K. developed the statistical model, prepared the population and covariate data. E.C. led the acquisition of all-cause mortality data. C.B. carried out the climate change attribution analysis and prepared the factual and counterfactual temperature data. D.X. extracted the results, created the plots and wrote the first draft of the paper and the supplement. All the authors contributed to modifying the paper and critically interpreting the results. All authors read and approved the final version for publication.

## Data sharing

The data used in this study were obtained from the Hellenic Statistical Authority (ELSTAT). Access to these data is subject to restrictions and was granted for the purposes of this research under specific authorization. The data are not publicly available but can be requested directly from ELSTAT, subject to their data access policies.

Air temperature at 2m for was retrieved from ERA5-land https://cds.climate.copernicus.eu/cdsapp#!/dataset/reanalysis-era5-land?tab=form.

## Data Availability

The data used in this study were obtained from the Hellenic Statistical Authority (ELSTAT). Access to these data is subject to restrictions and was granted for the purposes of this research under specific authorization. The data are not publicly available but can be requested directly from ELSTAT, subject to their data access policies.
Air temperature at 2m for was retrieved from ERA5-land https://cds.climate.copernicus.eu/cdsapp#!/dataset/reanalysis-era5-land?tab=form.

## Acknowledgements

G.K. is supported by an Imperial College Research Fellowship. This project has received funding from the European Union’s Horizon 2020 Research and Innovation program under the Marie Skłodowska-Curie Grant Agreement No 101008139 (EUREST-RISE).

