## Supplementary material for "Attributing heatwave mortality to human-induced climate change in Greece: a case-crossover and attribution analysis for 2000-2019": HeatwavesGreece_Supplementary.pdf

**Supplementary Tables**

**Supplementary Table 1. Descriptive statistics of population-weighted daily maximum temperature and relative humidity**

| NUTS3 region | Population-weighted daily maximum temperature (lag0) | | mean relative humidity (%, lag0) | |
|---|---|---|---|---|
| | Mean (SD) | Median [Min, Max] | Mean (SD) | Median [Min, Max] |
| N. ACHAIAS | 19.2 (8.01) | 18.2 [-2.21, 39.4] | 73.8 (12.3) | 76.2 [30.2, 96.1] |
| N. ANATOLIKIS ATTIKIS | 21.3 (8.33) | 20.6 [-1.12, 43.7] | 65.2 (14.1) | 66.3 [23.3, 93.8] |
| N. ARGOLIDAS | 21.1 (8.58) | 20.2 [-1.34, 43.0] | 66.8 (14.2) | 68.6 [24.8, 97.6] |
| N. ARKADIAS | 17.9 (8.46) | 17.1 [-4.53, 38.7] | 71.3 (15.0) | 73.5 [24.8, 98.3] |
| N. ARTAS | 18.8 (8.02) | 18.1 [-2.40, 39.9] | 76.6 (11.3) | 77.1 [32.0, 99.4] |
| N. ATHINON | 21.2 (8.37) | 20.3 [-0.967, 43.2] | 66.3 (14.0) | 67.7 [24.3, 94.9] |
| N. CHALKIDIKIS | 18.6 (7.78) | 18.2 [-3.38, 38.0] | 71.2 (10.8) | 71.3 [34.4, 95.6] |
| N. CHANION | 18.8 (6.13) | 18.2 [2.34, 35.2] | 71.4 (10.4) | 72.9 [22.8, 94.6] |
| N. CHIOU | 19.6 (7.22) | 18.9 [0.0571, 35.7] | 67.0 (11.1) | 67.1 [31.5, 93.6] |
| N. DODEKANISON | 20.6 (5.73) | 20.0 [4.19, 33.3] | 71.3 (8.90) | 72.8 [37.9, 92.1] |
| N. DRAMAS | 17.8 (8.52) | 17.5 [-4.86, 38.3] | 73.8 (11.4) | 74.4 [32.2, 99.1] |
| N. DYTIKIS ATTIKIS | 20.2 (8.47) | 19.3 [-2.10, 41.8] | 66.5 (14.3) | 67.7 [23.3, 96.8] |
| N. ETOLOAKARNANIAS | 19.6 (7.73) | 18.6 [-0.941, 39.9] | 74.3 (10.6) | 75.3 [35.1, 97.7] |
| N. EVROU | 18.9 (9.37) | 18.5 [-9.06, 40.7] | 69.0 (13.6) | 70.1 [25.7, 96.8] |
| N. EVRYTANIAS | 15.2 (8.30) | 15.0 [-9.27, 35.8] | 75.1 (14.1) | 76.3 [26.5, 99.4] |
| N. EVVIAS | 19.9 (7.76) | 19.5 [-1.14, 40.9] | 70.2 (11.8) | 70.6 [24.4, 96.4] |
| N. FLORINAS | 15.0 (9.49) | 14.9 [-14.0, 37.2] | 70.6 (15.2) | 71.4 [25.5, 99.6] |

| | | | | |
|---|---|---|---|---|
| N. FOKIDAS | 17.5 (8.20) | 16.9 [-5.30, 37.8] | 72.5 (12.6) | 74.4 [26.6, 96.5] |
| N. FTHIOTIDAS | 19.1 (8.82) | 18.5 [-4.79, 41.0] | 70.5 (14.9) | 72.0 [23.8, 98.2] |
| N. GREVENON | 16.8 (9.23) | 16.6 [-8.83, 38.4] | 67.2 (15.7) | 67.5 [24.9, 99.0] |
| N. ILIAS | 20.6 (7.37) | 19.4 [0.810, 39.8] | 74.4 (9.80) | 75.9 [31.3, 94.9] |
| N. IMATHIAS | 18.2 (8.93) | 17.9 [-7.69, 39.6] | 70.2 (13.8) | 70.8 [30.2, 98.2] |
| N. IOANNINON | 17.1 (8.72) | 16.5 [-5.67, 39.1] | 74.4 (13.6) | 75.1 [27.9, 99.8] |
| N. IRAKLIOU | 21.4 (6.99) | 20.9 [4.21, 40.2] | 71.2 (12.4) | 73.0 [20.3, 95.8] |
| N. KARDITSAS | 19.6 (9.15) | 19.0 [-4.14, 41.6] | 67.8 (15.4) | 69.3 [26.1, 97.9] |
| N. KASTORIAS | 15.9 (9.40) | 15.8 [-12.4, 37.5] | 70.1 (14.9) | 71.0 [23.4, 99.5] |
| N. KAVALAS | 17.3 (7.60) | 16.7 [-3.23, 34.0] | 73.2 (10.5) | 74.0 [34.1, 96.1] |
| N. KEFALLONIAS | 18.2 (5.57) | 17.5 [0.983, 29.8] | 74.1 (10.0) | 75.9 [35.1, 93.9] |
| N. KERKYRAS | 18.4 (5.89) | 17.6 [0.603, 32.7] | 73.8 (9.92) | 75.6 [28.6, 95.8] |
| N. KILKIS | 19.3 (9.28) | 18.9 [-7.50, 42.2] | 67.3 (14.4) | 67.3 [27.9, 97.8] |
| N. KORINTHOU | 19.1 (7.95) | 18.4 [-2.49, 38.4] | 67.4 (12.5) | 68.4 [28.0, 96.1] |
| N. KOZANIS | 17.0 (9.34) | 16.9 [-10.4, 38.7] | 68.0 (15.3) | 68.1 [23.7, 99.3] |
| N. KYKLADON | 18.9 (5.24) | 18.3 [4.33, 29.7] | 72.8 (8.06) | 73.6 [42.3, 94.0] |
| N. LAKONIAS | 20.3 (7.62) | 19.3 [0.696, 40.0] | 68.1 (12.2) | 69.3 [22.8, 94.6] |
| N. LARISAS | 20.3 (9.06) | 19.8 [-2.54, 43.2] | 65.9 (15.2) | 66.8 [19.0, 98.0] |
| N. LASITHIOU | 20.0 (6.25) | 19.4 [3.80, 35.2] | 68.5 (8.23) | 68.8 [30.0, 90.8] |
| N. LEFKADAS | 18.7 (5.97) | 18.0 [1.07, 34.0] | 75.8 (9.88) | 78.0 [33.5, 96.1] |
| N. LESVOU | 18.9 (7.33) | 18.0 [-0.527, 34.5] | 70.3 (9.78) | 70.3 [40.7, 95.0] |
| N. MAGNISIAS | 18.0 (7.78) | 17.8 [-2.94, 40.5] | 71.2 (12.0) | 71.6 [25.9, 97.9] |
| N. MESSINIAS | 19.8 (7.54) | 18.7 [0.741, 39.8] | 72.2 (10.6) | 73.2 [28.0, 95.9] |
| N. PELLAS | 18.5 (9.26) | 18.3 [-9.71, 40.9] | 69.8 (14.3) | 70.3 [27.7, 98.8] |
| N. PIERIAS | 18.2 (7.81) | 17.9 [-4.87, 37.4] | 72.2 (11.4) | 72.5 [31.8, 96.4] |
| N. PIREOS KE NISON | 21.6 (8.23) | 20.6 [0.305, 42.7] | 66.4 (12.8) | 67.4 [26.7, 95.5] |

| | | | | |
|---|---|---|---|---|
| N. PREVEZAS | 19.3 (7.45) | 18.4 [-0.378, 38.6] | 76.5 (9.93) | 77.6 [31.0, 97.4] |
| N. RETHYMNOU | 20.0 (6.95) | 19.4 [2.62, 38.3] | 69.6 (13.0) | 71.0 [19.2, 95.4] |
| N. RODOPIS | 18.4 (9.05) | 17.8 [-7.93, 37.6] | 70.7 (13.6) | 71.4 [31.1, 98.4] |
| N. SAMOU | 18.8 (5.81) | 18.3 [1.15, 31.5] | 70.7 (9.59) | 71.1 [43.2, 93.6] |
| N. SERRON | 19.4 (9.12) | 19.1 [-4.27, 41.1] | 68.7 (13.6) | 68.8 [30.1, 97.7] |
| N. THESPROTIAS | 18.6 (7.29) | 17.8 [-1.56, 38.5] | 75.6 (9.96) | 76.8 [30.2, 96.2] |
| N. THESSALONIKIS | 18.7 (8.50) | 18.4 [-6.08, 39.2] | 71.3 (11.8) | 71.4 [32.7, 97.1] |
| N. TRIKALON | 19.7 (9.35) | 19.2 [-4.84, 42.5] | 68.5 (15.5) | 69.5 [28.9, 98.9] |
| N. VIOTIAS | 20.0 (8.59) | 19.2 [-3.06, 42.0] | 68.6 (14.6) | 70.5 [23.8, 97.8] |
| N. XANTHIS | 17.3 (8.21) | 16.8 [-6.60, 35.5] | 72.9 (12.0) | 74.2 [34.5, 97.3] |
| N. ZAKYNTHOU | 18.9 (5.41) | 18.2 [2.36, 31.1] | 74.4 (9.36) | 75.8 [37.0, 93.9] |
| **Overall** | **19.7 (8.32)** | **19.1 [-14.0, 43.7]** | **69.4 (13.3)** | **70.7 [19.0, 99.8]** |

**Supplementary Table 2. Relative risk estimates for overall effect and effect modification by age and sex**

| | | HD1 | | HD2 | | HD3 | | HD4 | | HD5 | | HD6 | |
|---|---|---|---|---|---|---|---|---|---|---|---|---|---|
| **Sex** | **Age group** | **Unadjusted RR (95%CrI)** | **Adjusted RR (95%CrI)** | **Unadjusted RR (95%CrI)** | **Adjusted RR (95%CrI)** | **Unadjusted RR (95%CrI)** | **Adjusted RR (95%CrI)** | **Unadjusted RR (95%CrI)** | **Adjusted RR (95%CrI)** | **Unadjusted RR (95%CrI)** | **Adjusted RR (95%CrI)** | **Unadjusted RR (95%CrI)** | **Adjusted RR (95%CrI)** |
| Total | Total | 1.11 (1.11, 1.12) | 1.08 (1.07, 1.09) | 1.11 (1.1, 1.12) | 1.08 (1.06, 1.09) | 1.12 (1.11, 1.12) | 1.08 (1.07, 1.1) | 1.12 (1.12, 1.13) | 1.09 (1.07, 1.1) | 1.19 (1.17, 1.21) | 1.12 (1.1, 1.14) | 1.25 (1.21, 1.28) | 1.15 (1.11, 1.2) |
| | <75 | 1.05 (1.04, 1.06) | 1.04 (1.02, 1.05) | 1.05 (1.04, 1.06) | 1.03 (1.02, 1.05) | 1.06 (1.05, 1.07) | 1.05 (1.03, 1.06) | 1.07 (1.05, 1.08) | 1.05 (1.03, 1.07) | 1.12 (1.09, 1.15) | 1.09 (1.06, 1.12) | 1.16 (1.1, 1.22) | 1.12 (1.06, 1.19) |
| | 75-85 | 1.12 (1.11, 1.14) | 1.09 (1.07, 1.1) | 1.11 (1.1, 1.13) | 1.08 (1.06, 1.09) | 1.12 (1.11, 1.13) | 1.09 (1.07, 1.1) | 1.13 (1.12, 1.15) | 1.09 (1.07, 1.11) | 1.21 (1.18, 1.24) | 1.14 (1.11, 1.18) | 1.26 (1.2, 1.32) | 1.15 (1.09, 1.22) |
| | >85 | 1.16 (1.15, 1.17) | 1.12 (1.1, 1.13) | 1.15 (1.14, 1.17) | 1.11 (1.1, 1.13) | 1.16 (1.15, 1.18) | 1.12 (1.1, 1.14) | 1.17 (1.15, 1.18) | 1.11 (1.09, 1.13) | 1.23 (1.2, 1.26) | 1.15 (1.11, 1.18) | 1.31 (1.25, 1.36) | 1.2 (1.13, 1.27) |
| Males | Total | 1.08 (1.07, 1.09) | 1.06 (1.04, 1.07) | 1.08 (1.07, 1.09) | 1.06 (1.04, 1.07) | 1.09 (1.08, 1.1) | 1.06 (1.05, 1.08) | 1.09 (1.08, 1.1) | 1.07 (1.05, 1.08) | 1.15 (1.13, 1.17) | 1.11 (1.08, 1.13) | 1.19 (1.15, 1.24) | 1.13 (1.08, 1.19) |
| | <75 | 1.05 (1.03, 1.06) | 1.04 (1.02, 1.06) | 1.05 (1.04, 1.07) | 1.04 (1.02, 1.06) | 1.06 (1.04, 1.07) | 1.05 (1.03, 1.07) | 1.07 (1.05, 1.09) | 1.06 (1.03, 1.08) | 1.1 (1.07, 1.14) | 1.09 (1.04, 1.13) | 1.15 (1.08, 1.22) | 1.12 (1.04, 1.2) |
| | 75-85 | 1.09 (1.08, 1.11) | 1.06 (1.04, 1.09) | 1.08 (1.07, 1.1) | 1.06 (1.03, 1.08) | 1.09 (1.07, 1.11) | 1.06 (1.04, 1.09) | 1.09 (1.07, 1.12) | 1.07 (1.04, 1.09) | 1.16 (1.12, 1.2) | 1.13 (1.08, 1.17) | 1.18 (1.11, 1.26) | 1.13 (1.05, 1.22) |
| | >85 | 1.12 (1.1, 1.14) | 1.08 (1.06, 1.11) | 1.12 (1.1, 1.14) | 1.08 (1.06, 1.11) | 1.13 (1.11, 1.15) | 1.09 (1.06, 1.13) | 1.12 (1.1, 1.15) | 1.08 (1.04, 1.11) | 1.2 (1.15, 1.24) | 1.13 (1.08, 1.19) | 1.27 (1.18, 1.36) | 1.17 (1.07, 1.28) |

| | | | | | | | | | | | | | |
|---|---|---|---|---|---|---|---|---|---|---|---|---|---|
| Females | Total | 1.15 (1.14, 1.16) | 1.11 (1.09, 1.12) | 1.14 (1.13, 1.15) | 1.09 (1.08, 1.11) | 1.15 (1.14, 1.16) | 1.1 (1.09, 1.12) | 1.16 (1.15, 1.17) | 1.11 (1.09, 1.12) | 1.23 (1.21, 1.26) | 1.15 (1.12, 1.17) | 1.3 (1.26, 1.35) | 1.2 (1.15, 1.26) |
| | <75 | 1.06 (1.04, 1.08) | 1.03 (1, 1.06) | 1.04 (1.02, 1.06) | 1.01 (0.97, 1.04) | 1.07 (1.04, 1.09) | 1.04 (1.01, 1.08) | 1.07 (1.04, 1.1) | 1.04 (1.01, 1.08) | 1.14 (1.09, 1.19) | 1.1 (1.04, 1.15) | 1.18 (1.08, 1.28) | 1.12 (1, 1.23) |
| | 75-85 | 1.16 (1.14, 1.18) | 1.11 (1.09, 1.13) | 1.15 (1.13, 1.16) | 1.1 (1.08, 1.12) | 1.16 (1.14, 1.18) | 1.11 (1.09, 1.13) | 1.17 (1.15, 1.2) | 1.12 (1.09, 1.15) | 1.26 (1.22, 1.3) | 1.18 (1.13, 1.22) | 1.34 (1.26, 1.42) | 1.24 (1.15, 1.33) |
| | >85 | 1.19 (1.17, 1.2) | 1.14 (1.12, 1.16) | 1.18 (1.16, 1.19) | 1.13 (1.11, 1.15) | 1.18 (1.16, 1.2) | 1.13 (1.11, 1.15) | 1.2 (1.17, 1.22) | 1.13 (1.11, 1.16) | 1.26 (1.22, 1.29) | 1.16 (1.12, 1.2) | 1.33 (1.26, 1.41) | 1.2 (1.12, 1.29) |

### Supplementary Figures

**Supplementary Figure 1. Annual heatwave intensity and frequency under each definition.**

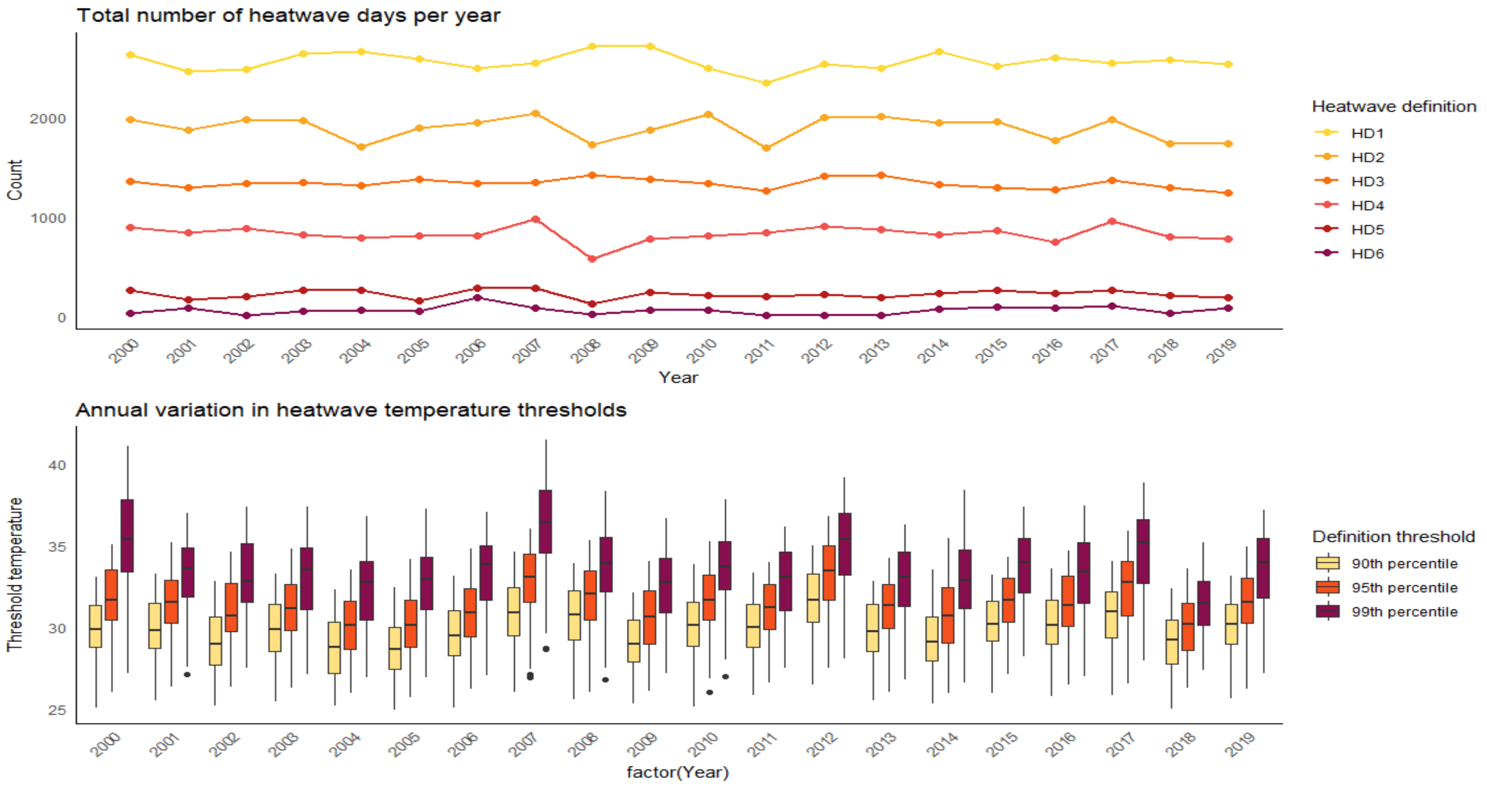

**Supplementary Figure 2. Total number of heatwave days under each definition over the study period in each NUTS3 region.**

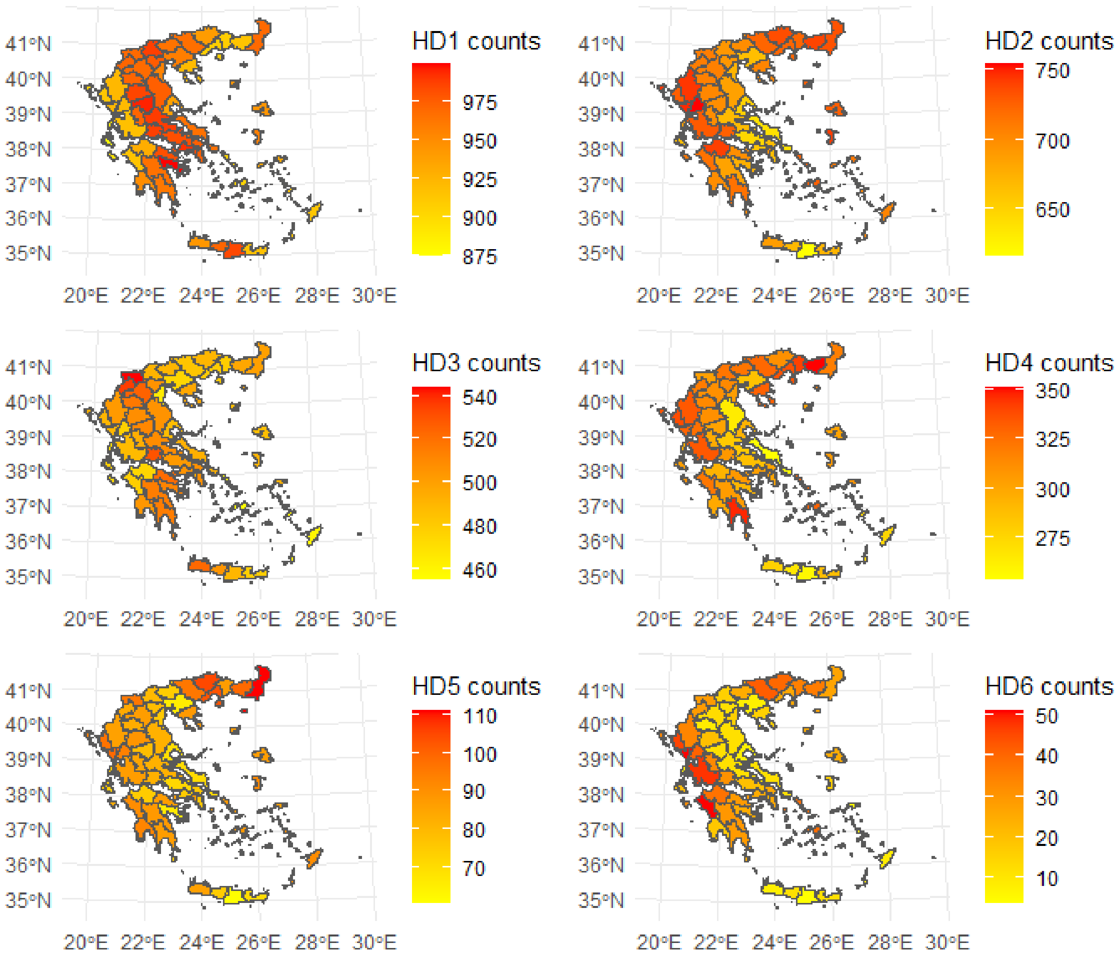

### Supplementary Texts

#### Supplementary Methods Section 1: Justification of population-weighted temperature

directly averaging the temperature across NUTS3 regions does not adequately represent the actual exposure experienced by the populations. This limitation arises from two aspects. First, temperature is retrieved at gridded level, while the mortality data are aggregated at NUTS3 region level, which causes spatial misalignment. Moreover, populations are not evenly distributed across space. Urban areas are usually densely populated, while some areas such as forests or mountains may be sparsely populated or uninhabited. For each NUTS3 region, we calculated its maximum daily temperature as a population-weighted average of the grid cell temperatures, using the same year's gridded population data from WorldPop at 1 km geographical resolution (www.worldpop.org).

$$T_{weighted} = \frac{\sum T_{grid} P_{grid}}{\sum P_{grid}}$$

#### Supplementary Methods Section 2: Counterfactual temperatures

The change in heatwave temperatures in each region associated with each degree of global warming was estimated using the probabilistic attribution protocol established by World Weather Attribution (1,2). We characterise heatwave temperatures in each region using Tx3x, the average of the daily maximum temperatures during the hottest three-day period in each year. A nonstationary Generalised Extreme Value (GEV) distribution is fitted to this time series of annual maxima, in which the location parameter of the distribution is assumed to shift linearly with global mean surface temperature (GMST), while the scale and shape parameters remain constant, so that

$$Tx3x\ GEV(\mu, \sigma | \mu_0, \sigma_0, \xi, \alpha, T) \text{ with } \mu = \mu_0 + \alpha T \text{ and } \sigma = \sigma_0,$$

Where $\mu, \sigma, \xi$ are the location, scale and shape parameters, $T$ is the GMST, and $\alpha$ is the change in local temperature per degree of GMST increase. To remove interannual variability, the GMST covariate is first smoothed using a 4-year running window. The parameters of this statistical model are obtained via maximum likelihood estimation. 95% confidence intervals are obtained by bootstrapping, using a sample size of 1000.

This procedure is repeated for two different observation-based datasets - E-Obs, which uses interpolated station data (3), and ERA5-land, a forecast-based reanalysis product(4) - and for 72 regional climate model runs from the Euro-CORDEX ensemble (5,6). The estimated $\alpha$ parameters from all of these data sources are then combined into a single, overarching estimate of the attributable change in local temperatures per degree of global warming, along with synthesised 95% confidence intervals, following the procedure described previously (7). This is done by first computing the weighted average of the estimates from the two observational datasets and, independently, the weighted average of the estimates from the 72 climate models; and then by calculating the weighted average of the two.

Having estimated the change in local heatwave temperatures per degree of global warming in this way, we calculate how much warmer each year was than a preindustrial climate with no or minimal human-caused warming (assumed to be 1.3°C cooler than 2025's mean temperature, per the Global Warming Index(8)). This temperature offset is multiplied by the alpha parameter and the estimated upper and lower bounds for each region to obtain an estimate and interval for local warming attributable to climate change. Finally, we subtract this attributable warming from the daily maximum temperatures in the ERA5-land dataset to obtain a counterfactual time series, along with upper and lower bounds. These three counterfactual time series are used as inputs to the epidemiological model, along with the factual time series (daily maximum temperatures averaged over each region from ERA5-land).

**Supplementary Methods Section 3: Model specifications**

We modelled the effect of heatwave by specifying Bayesian hierarchical conditional Poisson models, with a fixed effect on the event/non-event day grouping. This has been found to provide a flexible alternative to conditional logistic regression (9). Briefly, let $Y_{ijkl}$ taking the value 1 if it is an event and 0 otherwise on the $i$-th summer day, $j$-th NUTS3 region, $k$-th year and $l$-th event/non-event grouping. We have run the models for the different age-sex groups separately; therefore, for simplicity we omit an age-sex index from the notation:

$$Y_{ijkl} \text{ Poisson}(m_{ijkl})$$

$$\log(m_{ijkl}) = (b_1 + w_j + v_k) * X_{ijk} + b_2 Z_{ijk} + b_3 Z_{ik} + \phi_l$$

where $b_1$ is the average heatwave effect across the regions and years, $w_j$ is a Gaussian autoregressive prior that accounts for spatial dependence(10), allowing the average heatwave effect to take different values in the different NUTS3 regions. The random effect $v_k$ is a random walk of order 2 that models temporal autocorrelation, allowing the average heatwave effect to take different values across the different years. The term $b_2$ accounts for relative humidity, the term $b_3$ for national holidays and $\phi_l$ is a fixed effect per event/non-event grouping. We accounted for national holidays by including a binary variable, with 1 indicating national holiday and otherwise 0.

We report median and 95% Credible Intervals (CrI) of the posterior of the relative heatwave mortality risk by age, sex, heatwave definition, time and space.

**Supplementary Methods Section 4: Death attribution**

Omitting subscripts for simplicity, let $\bar{D}$ be the mean number of deaths during a heatwave day, *M* the number of heatwave days and RR the relative heatwave risk, then:

$$AN = \frac{RR - 1}{RR} * (M * \bar{D})$$

Let $AN_f$ denote the number of deaths in the *factual* scenario (based on the *observed* number of heatwave days $M_f$), and $AN_c$ the number of deaths in the counterfactual scenario without human-induced climate change (where the number of heatwave days $M_c$ is calculated using the counterfactual temperature time series). We report the median and 95% CrI for both the number and proportion of heatwave-related deaths attributable to climate change, defined respectively as $AN_f - AN_c$, and $(AN_f - AN_c)/AN_f$.

The heatwave-related relative mortality risks were estimated using thresholds derived from the factual temperature series to ensure consistency in the relative risks.

2. Philip S, Kew S, Van Oldenborgh GJ *et al.* A protocol for probabilistic extreme event attribution

analyses. *Adv Stat Clim Meteorol Oceanogr* 2020;**6**(2):177–203. https://doi.org/10.5194/ascmo-6-177-2020.
